# Precision Management of Fludrocortisone-Related Hypertension Risk in Congenital Adrenal Hyperplasia: A Machine Learning Approach to Personalized Dosing

**DOI:** 10.64898/2026.07.22.26358644

**Authors:** Shengkai Du, Zhaomin Chen, Gaohui Zhu, Taoting Li, Weiting Deng, Wenqing Ji, Yilu Yuan, Yansong Ba, Xinyu Wang, Rong Li

**Affiliations:** Chongqing Key Laboratory of Child Rare Diseases in Infection and Immunity, Department of Endocrine and Metabolic Diseases, Children’s Hospital of Chongqing Medical University, National Clinical Research Center for Children and Adolescents’ Health and Diseases, Chongqing 400016, P. R. China; College of Artificial Intelligence Medicine, Chongqing Medical University, 400016, Chongqing, China; Department of human resources, The First Affiliated Hospital of Chongqing Medical University, 400016, Chongqing, China; Lab Teaching & Management Center, Chongqing Medical University, Chongqing 400016, P. R. China; Laboratory of Tissue and Cell Biology, Experimental Teaching Center, Chongqing Medical University, Chongqing 400016, P. R. China

**Keywords:** Congenital adrenal hyperplasia, 21-hydroxylase deficiency, fludrocortisone, hypertension, machine learning, generalized linear mixed models

## Abstract

Congenital adrenal hyperplasia (CAH) is a rare inherited disorder requiring lifelong hormone replacement therapy. Excessive hormone replacement poses a significant risk for long-term complications, such as hypertension; however, quantitative approaches for optimizing dosing remain underdeveloped. This study aimed to identify factors associated with hypertension in patients with CAH and to develop a predictive model to support longitudinal fludrocortisone dose adjustment in pediatric patients who were already receiving mineralocorticoid replacement. We first employed generalized linear mixed models (GLMM) to evaluate the relationships among therapeutic agents, biochemical markers, and hypertension. Our results indicated a significant positive association between the dose of fludrocortisone (FC) and diastolic hypertension, whereas no such association was observed for the dose of hydrocortisone (HC). Using expert-curated data, we subsequently constructed multiple predictive models, —including CatBoost, XGBoost, and LightGBM, —to enable individualized adjustment of FC dosage. All models were evaluated on an independent test set, with CatBoost, XGBoost, and LightGBM demonstrating comparably strong performance (R²: 0.75–0.77). Subgroup analyses revealed that predictive accuracy was highest in children aged 0–2 years, where the top-performing model achieved a mean ideal prediction rate of 59.6%. This study not only confirms the significant link between FC dosing and hypertension in CAH patients but also provides a machine learning–based decision-support tool to assist individualized longitudinal dose adjustment. The model shows promise as a clinical decision-support instrument to facilitate personalized and precise management of CAH therapy.

## 1. Introduction

Classic congenital adrenal hyperplasia (CAH) is an autosomal recessive disorder caused by variants in key enzymes involved in adrenal steroidogenesis. The most prevalent form is 21-hydroxylase deficiency (21-OHD), where impaired enzyme activity leads to impaired glucocorticoid synthesis with or without mineralocorticoid deficiency. This deficiency causes compensatory over-secretion of adrenocorticotropic hormone (ACTH), adrenal cortex hyperplasia, and pathological accumulation of androgen and steroid precursors. This condition ultimately manifests as salt-wasting crises, ambiguous genitalia in 46,XX newborns, and other clinical features consistent with adrenal insufficiency (Auer et al., 2023). Long-term replacement therapy is essential, with the standard regimen consisting of glucocorticoid supplementation combined with mineralocorticoid replacement. Hydrocortisone (HC) is the glucocorticoid of choice for replacement therapy, particularly in pediatric patients before they reach adult height, while fludrocortisone (FC) serves as the primary mineralocorticoid. The therapeutic goal is to prevent both hyperandrogenism and adrenal crises while minimizing treatment-related adverse effects (Speiser et al., 2018).

Among various dose-related complications, hypertension has particularly received clinical attention due to its high prevalence and potential long-term cardiovascular consequences (Barbot et al., 2022). Multiple studies have reported a higher incidence of hypertension in CAH patients compared to the general population, with an elevated risk persisting across age groups (Bonfig and Schwarz, 2014; Falhammar et al., 2015; Improda et al., 2019). A longitudinal cohort study by Bao et al. demonstrated that blood pressure levels in infancy and early childhood are positively correlated with adult blood pressure values. This finding suggests that early-life blood pressure dysregulation may predispose individuals to cardiovascular disease in adulthood (Bao et al., 1995). Cardiovascular diseases remain the leading cause of mortality and morbidity worldwide, posing a major threat to global public health. In 2022 alone, an estimated 19.8 million deaths were attributed to cardiovascular causes, accounting for approximately 32% of total global mortality. Thus, precise titration of replacement therapy to prevent hypertension in CAH patients is an urgent clinical priority.

However, current clinical practice relies heavily on dynamic monitoring of biochemical markers and clinical signs for empirical dose adjustment (Auer et al., 2023). This reactive approach is inherently limited by individual variability in dosing requirements and age-dependent physiological changes, making proactive and precise dose optimization challenging.

The rapid development of machine learning offers a promising avenue to address this clinical dilemma. Compared with traditional statistical methods, machine learning can automatically capture non-linear relationships and interactions among variables, providing distinct advantages in modeling complex medical data. It is particularly well-suited for handling high-dimensional, multifactorial clinical datasets and shows great potential in individualized follow-up management. Therefore, this study aimed to investigate factors associated with hypertension in pediatric patients with classic CAH and to develop a predictive model to support longitudinal fludrocortisone dose adjustment during follow-up. We first employ generalized linear mixed models (GLMM) to identify intervention factors significantly associated with hypertension status. We then developed machine-learning models to predict follow-up fludrocortisone dose among patients already receiving mineralocorticoid replacement, thereby offering a novel strategy for improving treatment outcomes and reducing the risk of adverse events in CAH patients.

## 2. Materials and Methods

### 2.1. Study Population

This study utilized a large-scale, real-world longitudinal cohort, retrospectively including 1,403 CAH patients with a total of 5,960 clinical visits at the Children’s Hospital of Chongqing Medical University from 2014 to 2024. To ensure the cohort’s homogeneity and alignment with the research objectives, stringent exclusion criteria were applied during data cleaning. The following records were removed:

(1) Clinically unstable episodes, which included visits documenting adrenal crises and records from newly diagnosed patients with less than three months of medication exposure (Auer et al., 2023); (2) Patients with nonclassic CAH (NCCAH) or non-21-hydroxylase deficiency; (3) Visits involving patients older than 12 years (Lawrence et al., 2022; Kim et al., 2017), which affected 3.6% of records; (4) Individuals not receiving standard HC and FC replacement therapy; and (5) Records with biologically implausible data (e.g., height >200 cm or weight >100 kg).

The detailed screening workflow is presented in Figure 1. The study protocol was approved by the Ethics Committee of the Children’s Hospital of Chongqing Medical University, with a waiver of informed consent granted in accordance with the ethical approval documentation.

**Figure 1.**
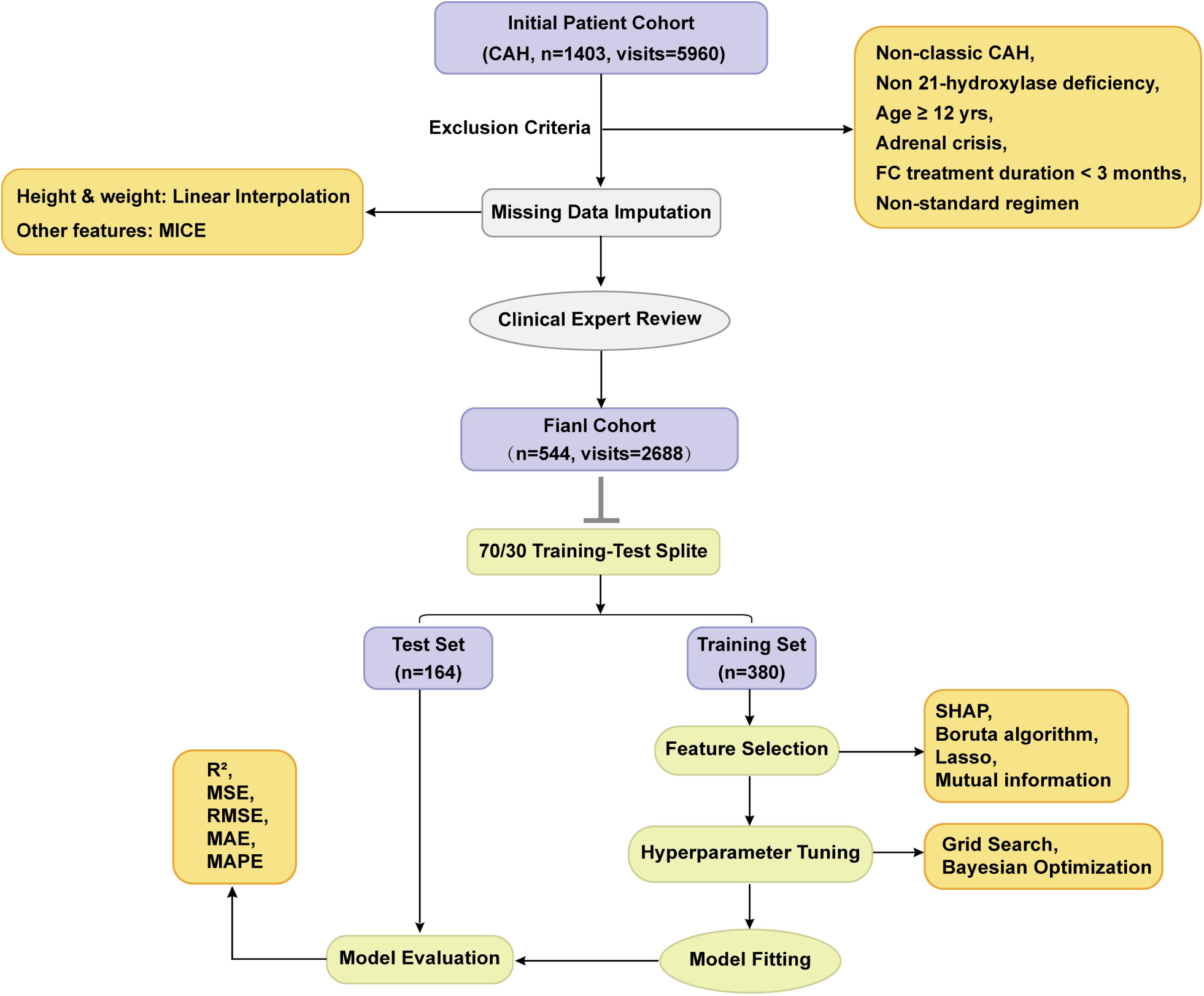
Flowchart of dose prediction algorithm development and evaluation. CAH, congenital adrenal hyperplasia; MICE, multiple imputation by chained equations; SHAP, shapley additive explanations.

### 2.2. Data Extraction and Preprocessing

The extracted variables included: (1) laboratory results (17-hydroxyprogesterone, plasma renin concentration (PRC), sodium, etc.); (2) clinical measurements (height, weight, blood pressure, etc.); and (3) medication records (fludrocortisone, hydrocortisone, salt supplementation). All data were sourced from the hospital’s electronic medical records, laboratory information, and pharmacy management systems, linked via unique patient and visit identifiers. FC and HC doses were converted to total daily doses and normalized to body surface area. The “previous dose”, defined as the medication dosage from a patient’s immediately preceding visit, was included to represent the longitudinal treatment baseline used in follow-up dose adjustment. Height standard deviation scores (Height-SDS) and body mass index standard deviation scores (BMI-SDS) were calculated according to the Chinese Growth Reference Standards for Children and Adolescents Aged 0–18 Years (Li, n.d.). Blood pressure standard deviation scores (BP-SDS) were computed using the R package pedbp with age-stratified references: Gemelli et al.’s longitudinal data (1990) for infants under 1 year (Gemelli et al., 1990) and the 2017 clinical practice guidelines by Flynn et al. for children aged 1–12 years (Flynn et al., 2017).

### 2.3. Outlier and Missing Data Handling

Outliers were identified using Tukey’s method (values below Q1 - 3×IQR or above Q3 + 3×IQR). For variables demonstrating significant age-dependent variation (e.g., medication doses, PRC), outlier detection was performed within age-stratified groups (0–2, 2–5, 5–8, >8 years). Identified outliers were treated as missing values (NA) and were subsequently imputed.

A hierarchical hybrid approach was employed for missing data: records lacking age or sex information were excluded. Missing height and weight data were addressed via linear interpolation. For all other variables, including medication doses, biochemical markers, and blood pressure, multiple imputation by chained equations (MICE) was implemented, generating 10 complete imputed datasets (Buuren and Groothuis-Oudshoorn, 2011). These datasets were utilized exclusively for GLMM analysis and prediction model development, with results pooled across the imputed datasets using Rubin’s rules (Rubin, 1987).

### 2.4. Statistical Analysis

All statistical analyses were performed using R (version 4.5.1) and/or Python (version 3.12.4)

#### 2.4.1. Descriptive Statistics

The normality of continuous variables was assessed using the Shapiro-Wilk test. As most variables exhibited skewed distributions, data are summarized as median and interquartile range (IQR). Categorical variables are presented as frequencies and percentages. Comprehensive baseline characteristics of the study population are detailed in Table 1.

**Table 1.** Baseline Characteristics of the Study Cohort.

| Variables | Median (IQR) | Missing Percentage (%) |
| --- | --- | --- |
| Number of patients | 724 | 0 |
| Number of visits | 4324 | 0 |
| Age | 3.3 (1.6 to 6.2) | 0 |
| Subtype, n(%) |  | 0 |
| SW | 277 (38.3%) |  |

**Table 1.** Baseline Characteristics of the Study Cohort.
| Variables | Median (IQR) | Missing Percentage (%) |
| --- | --- | --- |
| SV | 447 (61.7%) |  |
| Sex, n(%) |  | 0 |
| Female, n (%) | 329 (45.4%) |  |
| Male, n (%) | 395 (54.6%) |  |
| Height SDS | -0.82 (-1.60 to 0.03) | 0.2 |
| BMI SDS | 0.51 (-0.16 to 1.30) | 0.2 |
| Blood Pressure (mmHg) |  | 26.6 |
| Systolic | 101 (94 to 109) |  |
| Diastolic | 59 (53 to 65) |  |
| Blood Pressure SDS |  | 26.6 |
| Systolic | 0.78 (0.19 to 1.46) |  |
| Diastolic | 0.64 (0.09 to 1.20) |  |
| Hypertension (%) | 26.7 | 26.6 |
| Fludrocortisone ($\mu\text{g}/\text{m}^2/\text{d}$) | 68.6 (44.3 to 106.6) | 12.8 |
| Hydrocortisone ($\text{mg}/\text{m}^2/\text{d}$) | 12.1 (9.6 to 15.3) | 10.9 |
| 17-OHP (ng/mL) | 8.2 (1.7 to 26.6) | 29.9 |
| PRC ($\mu\text{IU}/\text{mL}$) | 33.6 (10.1 to 85.9) | 20.9 |
| Sodium (mmol/L) | 140.4 (139.0 to 141.9) | 0.66 |
| Potassium (mmol/L) | 4.5 (4.2 to 4.8) | 0.66 |

SDS, standard deviation scores. Data are presented as median (IQR) for continuous variables and n (%) for categorical variables.

**Table 2.** Univariable analysis of factors associated with systolic and diastolic hypertension.

| Parameter | Diastolic Hypertension |  | P value | Systolic Hypertension |  | P value |
| --- | --- | --- | --- | --- | --- | --- |
|  | No Hypertension | Diastolic Hypertension |  | No Hypertension | Systolic Hypertension |  |
| Fludrocortison e ($\mu\text{g}/\text{m}^2/\text{d}$) | 61.5 (40.7 to 92.1) | 97.0 (67.2 to 130.8) | < 0.001 | 61.5 (39.2 to 95.0) | 65.3 (38.8 to 105.3) | 0.002 |
| Hydrocortisone (mg/m<sup>2</sup>/d) | 12.5 (10.0 to 16.0) | 11.7 (9.1 to 14.7) | < 0.001 | 12.5 (10.0 to 16.0) | 12.6 (9.8 to 16.7) | 0.8 |
| 17-OHP (ng/mL) | 8.8 (2.1 to 27.6) | 6.6 (1.8 to 20.7) | 0.04 | 8.8 (2.1 to 27.6) | 9.1 (1.7 to 29.6) | 0.5 |
| PRC ($\mu\text{IU}/\text{mL}$) | 37.6 (14.2 to 86.0) | 26.3 (3.7 to 76.1) | < 0.001 | 37.6 (14.2 to 86.0) | 34.1 (8.0 to 84.8) | 0.005 |
| Sodium (mmol/L) | 140.2 (139.0 to 141.6) | 140.4 (139.1 to 142.0) | 0.01 | 140.2 (139.0 to 141.6) | 140.6 (139.1 to 142.1) | < 0.001 |
| Potassium (mmol/L) | 4.5 (4.2 to 4.8) | 4.6 (4.2 to 4.9) | 0.004 | 4.5 (4.2 to 4.8) | 4.4 (4.1 to 4.7) | 0.002 |

<sup>†</sup> Mann-Whitney U test. P<0.05 is considered significant.

#### 2.4.2. Analysis of Factors Associated with Hypertension

Hypertension status was defined as either a single-visit systolic or diastolic BP-SDS > 1.645 (corresponding to the 95th percentile). In the text below, this will be uniformly referred to as ’hypertension’. To identify independent factors associated with hypertension, we first conducted univariable analyses using Mann-Whitney U tests with Bonferroni correction to compare distributions between hypertensive and non-hypertensive groups. Subsequently, multivariable analysis was performed using GLMM. This model integrates the flexibility of generalized linear models for non-normal, binary outcomes with the capacity of mixed-effects models to account for non-independence among repeated measurements via patient-specific random intercepts, enabling more precise parameter estimation (Bates et al., 2015).

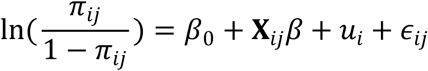

In these models, hypertension (yes/no) served as the binary outcome. Fixed-effects covariates included age, sex, height, weight, disease subtype and other medication use to control for potential confounding. Key predictors, including FC dose, HC dose, PRC, 17-hydroxyprogesterone, sodium, and potassium, were incorporated to evaluate their relationships with hypertension. Because all continuous clinical and biochemical variables exhibited non-normal distributions, they were first log-transformed and then standardized before modeling. Additionally, to evaluate the quantitative association between FC dose and PRC, we constructed a linear mixed-effects model (LMM) with PRC as the dependent variable and FC dose as the primary predictor. The model was adjusted for aforementioned potential confounders.

Sensitivity analyses were further performed under different delta-adjustment assumptions to assess the robustness of these associations (Gachau et al., 2020).

### 2.5. Development of A Fludrocortisone Dose Prediction Model

Building upon GLMM findings, we developed machine-learning models intended as predictive decision-support tools.

#### 2.5.1. Dataset Construction and Feature Engineering

Pediatric endocrinologists reviewed all visits, selecting those that appropriately reflected treatment regimens based on clinical signs and biochemical markers documented during subsequent follow-up visits. Two independent experts performed blinded assessments, and any disagreements were resolved by a third arbitrator. The FC dose from expert-validated visits served as the continuous target variable for prediction.

The dataset then underwent another MICE imputation cycle, excluding the target variable (current FC dose) and the “previous dose” feature to prevent data leakage. Imputation quality was evaluated by comparing kernel density plots of imputed versus original distributions and calculating Kullback-Leibler divergence between the imputed datasets. The imputed set that best matched the original data distribution was selected for modeling (Supplementary Figure S1). Prior to modeling, Pearson correlation analysis and variance inflation factor (VIF) assessment confirmed the absence of severe multicollinearity.

#### 2.5.2. Model Building

The dataset was grouped by patient ID, and these groups were then randomly divided into training and test sets (70:30) to prevent information leakage. Given the interrelated adjustment of FC and HC dosing, monitoring indicators for both medications were included in the initial candidate feature set to capture potential interactions (Liu et al., 2013). To refine the feature set, we applied four feature selection algorithms, including Boruta, mutual information, Lasso, and Random Forest-based SHAP, on the training set. Features were selected based on consensus voting across these methods, supplemented by input from clinical experts.

Six machine learning models, including CatBoost, XGBoost, LightGBM, Support Vector Regression (SVR), K-Nearest Neighbors (KNN), and Linear Regression (LR), were trained using the selected features. Model stability was ensured via 10-fold cross-validation, with hyperparameter optimization performed using Bayesian optimization and grid search. Predictive performance was evaluated on the independent test set using root mean square error (RMSE), mean absolute error (MAE), mean absolute percentage error (MAPE), and the coefficient of determination (R²). For robust estimation, bootstrap resampling (100 iterations) was used to compute the median and 95% confidence interval (95% CI) for each metric (Li et al., 2021).

To evaluate the incremental predictive contribution of previous FC dose, we performed a comparative feature ablation and baseline analysis involving three models: a full model including previous dose and all selected predictors, a no-previous-dose model excluding the previous-dose feature, and a previous-dose-only baseline model using previous dose as the sole predictor. For comparability, the same best-performing algorithm identified in the full-model analysis was used for the no-previous-dose and previous-dose-only models. All three models used the same patient-level training-test split, cross-validation framework, hyperparameter optimization strategy, and evaluation metrics.

#### 2.5.3. Model Comparison and Interpretability

The coefficient of determination (R²) served as the primary metric for model comparison. To evaluate the stability of the final selected model across different patient subgroups, the test cohort was stratified into three age groups (0–2, 2–8, and 8–12 years). Prediction accuracy within these subgroups was quantified using error margins: predictions within ±20% of the true value were classified as ’Ideal,’ those exceeding 120% as ’Over,’ and those below 80% as ’Under’ (Steiner et al., 2021). Overall differences across groups and models were assessed using the Friedman test, followed by post-hoc pairwise comparisons conducted using the Wilcoxon signed-rank test with Bonferroni correction.

To enhance model transparency and clinical trustworthiness, we applied shapley additive explanations (SHAP) to the best-performing model, elucidating the contribution of individual features to the final dose prediction. SHAP values were interpreted as measures of model contribution rather than causal effects.To further examine whether the contribution of previous FC dose varied according to current clinical and biochemical indicators, SHAP interaction values were calculated between previous dose and other core features. Variables were dichotomized into high- and low-level groups based on the median, and locally weighted scatterplot smoothing (LOWESS) was applied to visualize trend patterns.

#### 2.5.4. Ethics Statement

This study was approved by the Institutional Review Board (IRB) of the Children’s Hospital of Chongqing Medical University (Approval No. [2023] Lun-Shen-Yan No. 165).

Written informed consent was not required for participation in this study in accordance with national legislation and institutional requirements.The study was conducted in accordance with the ethical principles of the World Medical Association (WMA) and the Council for International Organizations of Medical Sciences (CIOMS).

## 3. Results

### 3.1. Baseline Characteristics

This study initially enrolled 1,403 patients, with a total of 5,960 clinical visits. After excluding 1,636 visits that did not meet the inclusion criteria, the final analytical cohort comprised 724 patients with 4,324 visits. Among these patients, 45.4% were female, and 54.6% were male. Regarding CAH subtypes, 61.7% were classified as the simple virilizing (SV) form and 38.3% as the salt-wasting (SW) form. The median number of visits per patient was 5 (IQR: 3–8), and the median age at the time of visits was 3.3 years (IQR: 1.6–6.2).

Table 1 summarizes the baseline characteristics and the status of missing data for each variable. The overall prevalence of hypertension across all visits was 26.7%. Mineralocorticoid (fludrocortisone, FC) and glucocorticoid (hydrocortisone, HC) doses were considered key intervention variables potentially influencing this outcome. Mineralocorticoid replacement therapy was administered to 87.2% of patients, exclusively using FC, with a median dose of 68.6 μg/m²/d. All patients received glucocorticoid replacement therapy, with HC used in 99.1% of visits at a median dose of 12.1 mg/m²/d. Blood pressure data had a high proportion of missing values (26.6%), primarily concentrated among visits of children aged 0–1 year, where compliance with measurements was lower.

### 3.2. Factors Associated With Hypertension

The age-dependent trends for BP-SDS and medication doses are shown in Figure 2. The results indicate that the trend for DBP-SDS consistently declined with age, similar to the observed pattern for FC dosage. DBP-SDS was notably higher in younger patients and decreased as age increased, whereas SBP-SDS remained relatively stable across age groups. The median FC dose demonstrated a significant decline with age, while the median HC dose decreased during the 0–1 year period before slowly increasing thereafter.

**Figure 2.**
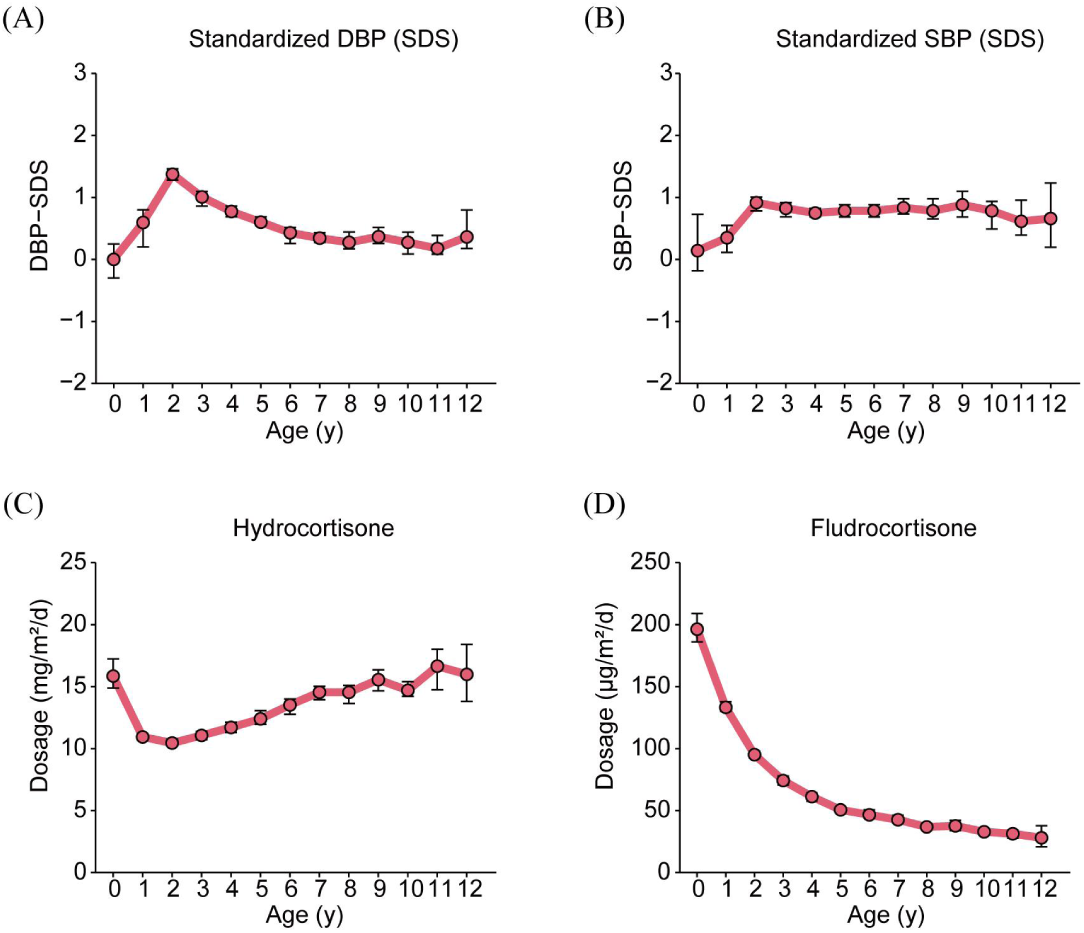
Line plots depict the longitudinal trends of **(A)** standardized systolic blood pressure (SBP-SDS), **(B)** standardized diastolic blood pressure (DBP-SDS), **(C)** hydrocortisone (HC) dosage, and **(D)** fludrocortisone (FC) dosage across ages 0 to 12 years. Each data point represents the median value aggregated from all patient visits within a ±6-month window around the corresponding age.

Next, we compared visits with documented hypertension to those without. Mann-Whitney U tests revealed that visits involving diastolic hypertension were associated with a higher median FC dose (95.2 μg/m²/d) and a lower median HC dose (12.7 mg/m²/d) compared to visits without hypertension. In the systolic hypertension group, only a higher FC dose (65.3 μg/m²/d) was observed, with no statistically significant difference found in HC dose. Additionally, lower PRC levels were noted in both hypertensive groups (Diastolic Hyper: 26.3 μIU/L; Systolic Hyper: 34.1 μIU/L). Although there were statistically significant differences in serum sodium and potassium levels between the groups, the absolute differences were minimal and of little clinical significance.

To identify factors independently associated with hypertension, we constructed GLMMs, with missing variables addressed with the hierarchical hybrid approach described earlier. The models were adjusted for age, height, weight, disease subtype, and concurrent medication use as potential confounders. A binary variable (0–1 year vs. 1–12 years) was included to account for potential methodological bias due to the switch in BP-SDS reference standards.

Results from the GLMM analysis (Figure 3) indicated that, after adjusting for confounders, a higher FC dose was significantly associated with an increased risk of diastolic hypertension. Sensitivity analyses demonstrated that the significant positive association between FC dose and diastolic hypertension remained consistent across different delta adjustment levels (Supplementary Figure S1). Conversely, the HC dose showed no significant association with either systolic or diastolic hypertension. The association between 17-hydroxyprogesterone and hypertension was also insignificant. On the other hand, lower PRC was independently associated with both diastolic and systolic hypertension, and lower potassium and higher sodium levels were associated with systolic hypertension.

**Figure 3.**
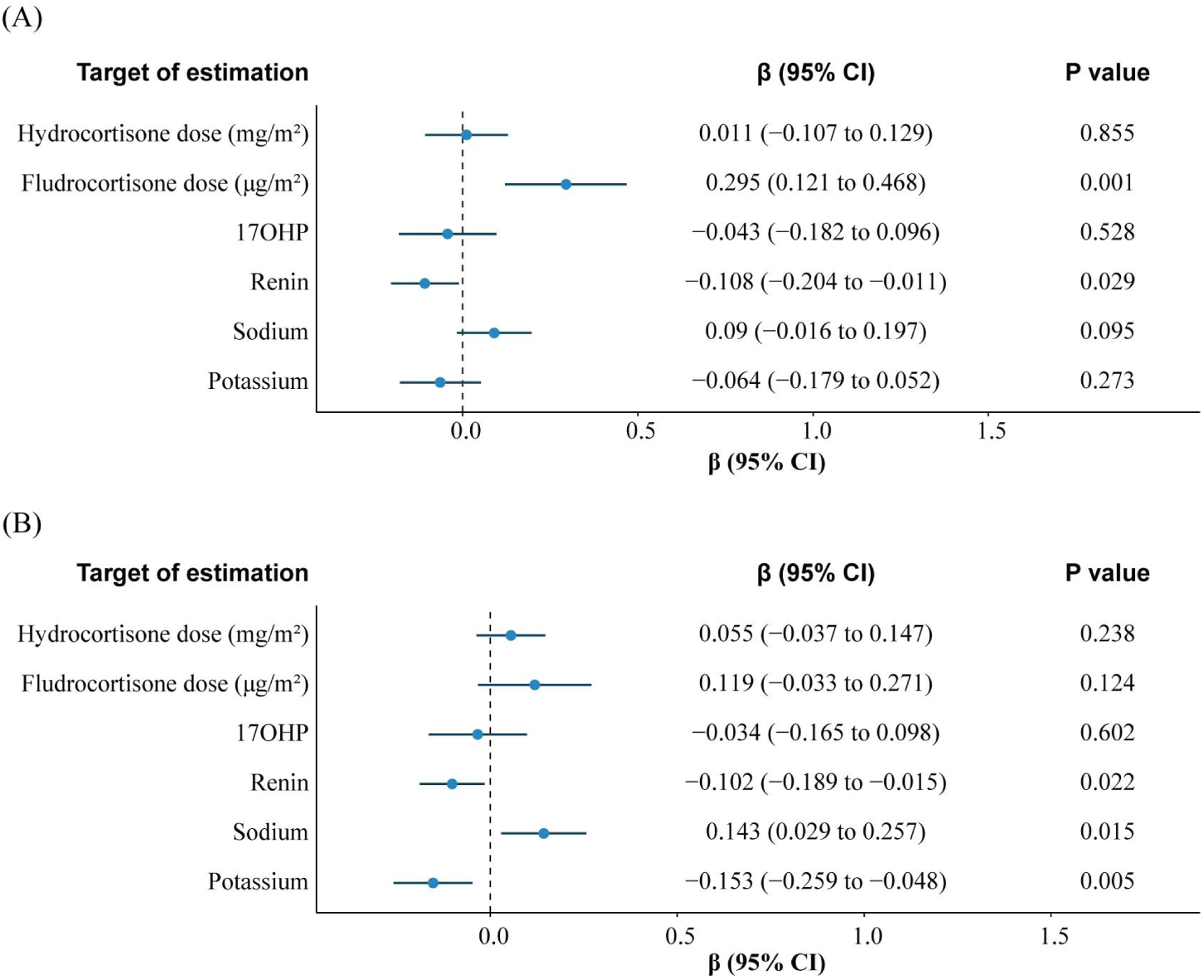
Forest plots display pooled β coefficients (95% CI) from generalized linear mixed models (GLMM). **(A)** Models with systolic hypertension as response variable. **(B)** Models with diastolic hypertension as response variable. An association is considered statistically non-significant if the 95% CI crosses the null value (β=0).

In the additional LMM, higher FC dose was significantly associated with lower PRC after adjustment for prespecified potential confounders (β = -0.1313, 95% CI: -0.2057 to -0.0569, P = 0.0011). Because both variables were log-transformed and standardized, to aid interpretation, we translated the standardized coefficient back to the log-log scale. The corresponding unstandardized log-log coefficient was approximately -0.301. This indicates that a 1% increase in FC dose was associated with an approximately 0.30% decrease in PRC. Sensitivity analyses showed that this inverse association remained consistent across different delta-adjustment levels (Supplementary Figure S2).

### 3.3. Key Feature Selection for Model Construction

Building upon the GLMM results, we proceeded with feature selection to define an optimal feature set for constructing machine learning models to predict FC dose. Seventeen clinical and biochemical features were initially considered. Four distinct methods, including Boruta algorithm, mutual information, Lasso regression, and Random Forest-based SHAP analysis, were employed to rank variable importance (Figure 4). Ultimately, seven variables, including previous dose, age, PRC, potassium, SBP-SDS, DBP-SDS, and Height-SDS, were consistently ranked highly by at least three methods (Figure 4). Among them, previous dose and age were identified as the most influential predictors.

**Figure 4.**
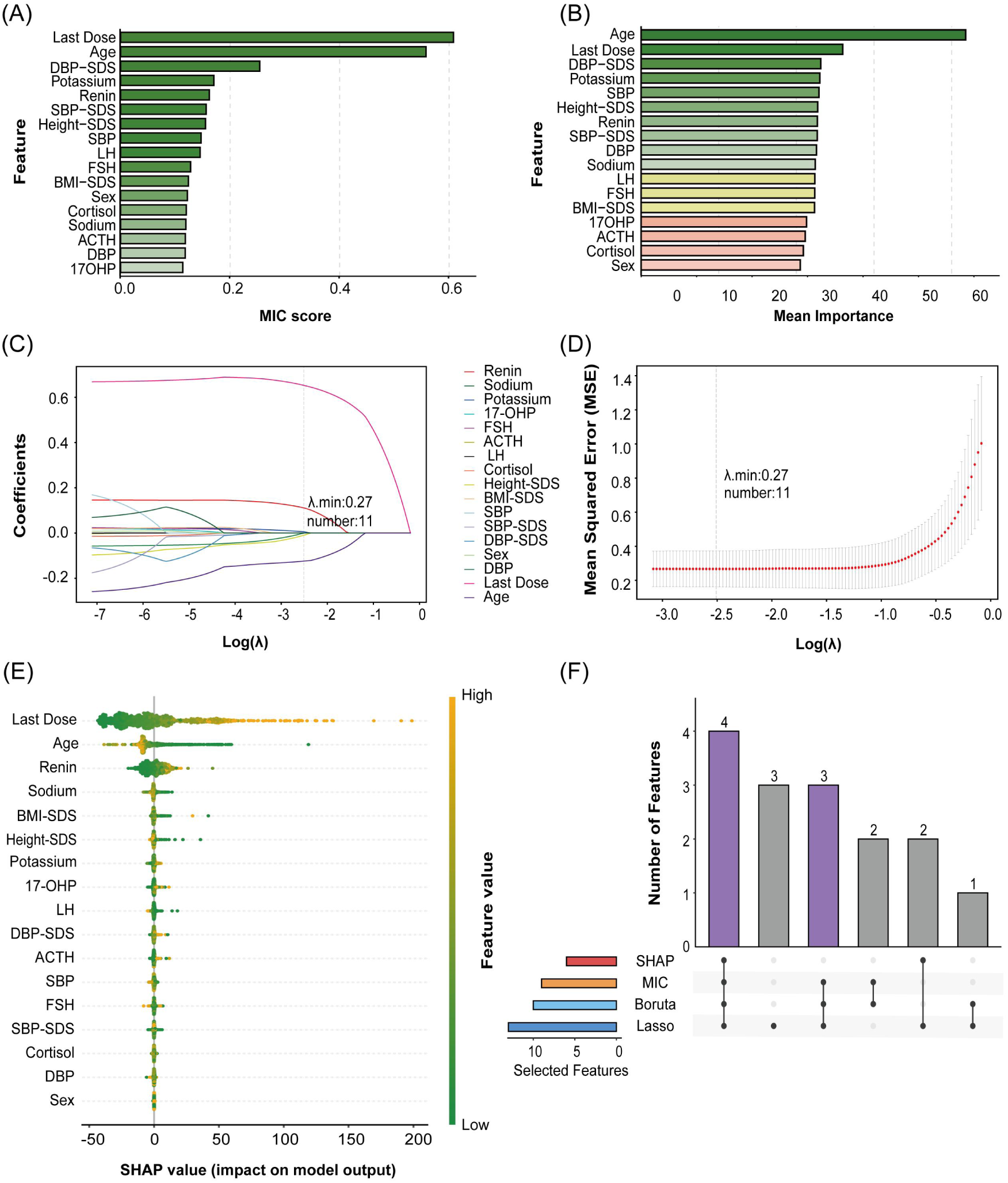
Feature Selection Using Four Distinct Algorithms. **(A)** Mutual information scores. **(B)** Boruta algorithm output: features are classified as Confirmed (green), Tentative (yellow), or Rejected (red). **(C)** Regularization path of Lasso regression, showing how coefficients shrink as the penalty (λ) increases. **(D)** Cross-validation curve for Lasso; λ.min denotes the value of λ at which the mean cross-validated error is minimized, with the corresponding number of non-zero features indicated. **(E)** Feature importance based on SHAP values from Random Forest model. **(F)** Upset plot illustrating the consensus of features selected by each algorithm. Features selected by at least three methods were retained for the final model. MIC, mutual information coefficient; SDS, standard deviation scores; LH, luteinizing hormone; FSH, follicle-stimulating hormone; ACTH, adrenocorticotropic hormone; 17-OHP, 17-hydroxyprogesterone; SBP, systolic blood pressure; DBP, diastolic blood pressure; SBP-SDS, systolic blood pressure standard deviation scores; DBP-SDS, diastolic blood pressure standard deviation scores.

### 3.4. Performance Comparison of Predictive Models

Model performance comparisons revealed that CatBoost and XGBoost achieved the best performance on the test set, both with a median R² of 0.77 (Table 3), indicating substantial ability to explain inter-individual variation in fludrocortisone dose. Scatter plots of predicted versus observed doses for each model in the test cohort are shown in Figure 5; the diagonal line represents perfect prediction, while points above or below this line indicate over- or under-estimation, respectively.

**Table 3.** The comparative performance of each algorithm is quantified by four metrics, reported as median (95% Cl) estimated from 1000 bootstrap replicates of the test set.

| Model | R<sup>2</sup> | MAE | RMSE | MAPE |
| --- | --- | --- | --- | --- |
| CatBoost | 0.77 (0.72-0.83) | 17.88 (16.60-19.69) | 29.25 (24.72-34.25) | 0.33 (0.29-0.38) |
| KNN | 0.67 (0.59-0.73) | 21.54 (20.00-23.45) | 35.05 (30.06-42.22) | 0.36 (0.36-0.43) |
| LightGBM | 0.75 (0.68-0.82) | 18.09 (16.56-20.51) | 29.97 (24.46-36.37) | 0.32 (0.28-0.40) |

**Table 3.** The comparative performance of each algorithm is quantified by four metrics, reported as median (95% CI) estimated from 1000 bootstrap replicates of the test set.
| Model | R<sup>2</sup> | MAE | RMSE | MAPE |
| --- | --- | --- | --- | --- |
| LR | 0.69 (0.61-0.80) | 20.10 (18.34-22.14) | 33.36 (27.49-39.70) | 0.34 (0.31-0.39) |
| SVR | 0.72 (0.63-0.78) | 18.35 (16.38-20.55) | 32.56 (27.61-40.41) | 0.29 (0.26-0.34) |
| XGBoost | 0.77 (0.71-0.82) | 17.95 (16.25-19.57) | 29.28 (25.03-33.77) | 0.32 (0.28-0.37) |

R<sup>2</sup>, coefficient of determination; MAE, mean absolute error; RMSE, root mean square error; MAPE, mean absolute percentage error.

**Figure 5.**
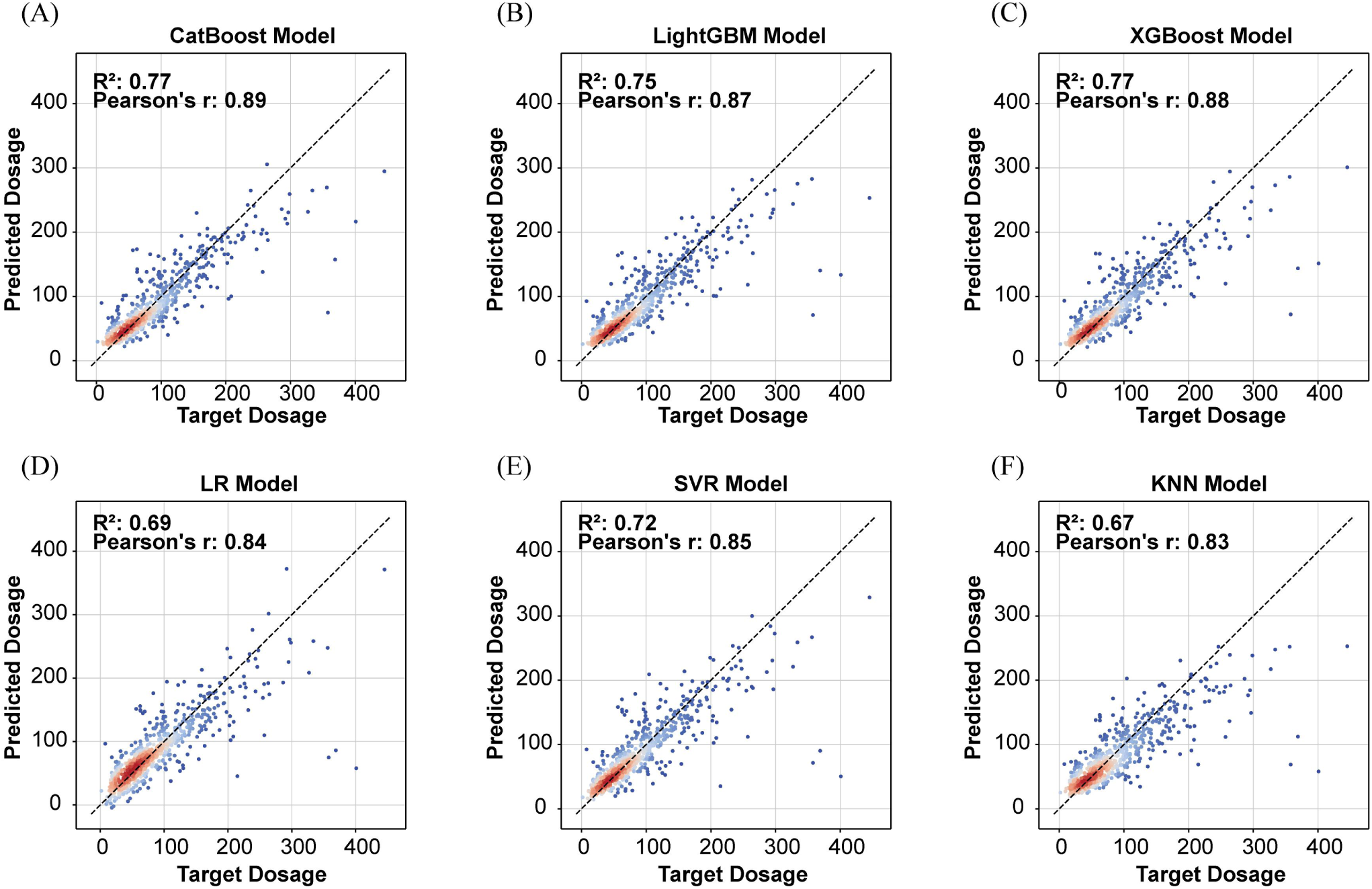
Scatter plots compare model predictions against the true values for each algorithm. The diagonal line represents the line of perfect agreement. Warmer colors indicate higher data point density. R² and Pearson’s correlation coefficient are reported for each model.

To statistically compare the performance differences between models, we performed the Friedman test followed by Bonferroni-corrected Wilcoxon signed-rank tests (α’ = 0.003), which indicated a significant overall difference among models (P < 0.001). However, no significant difference was found between CatBoost and XGBoost (P = 0.30), and the difference between XGBoost and LightGBM did not reach the corrected significance threshold (P = 0.009). Therefore, CatBoost, XGBoost, and LightGBM were selected as the top-performing models for further analyses.

To assess the contribution of previous FC dose, we conducted a three-model comparison using the CatBoost algorithm. The full model achieved the best predictive performance, with an median R² of 0.77. The no-previous-dose model retained moderate predictive performance, with an median R² of 0.59. The previous-dose-only baseline model achieved an median R² of 0.56 (Table S1).

We also evaluated model performance across different age groups by conducting separate analyses within three subgroups: 0–2 years, 2–8 years, and 8–12 years (Table S2).The mean ideal prediction percentage was highest in the 0–2 years subgroup (59.6%), significantly outperforming the 2–8 years (53.8%) and 8–12 years (43.2%) subgroups, suggesting superior predictive accuracy for younger patients. Regarding model comparisons within these subgroups, CatBoost slightly outperformed XGBoost and LightGBM in the 0–2 years subgroup, while LightGBM showed the best performance in the 2–8 years subgroup. In the 8–12 years subgroup, XGBoost and LightGBM showed comparable performance, both exceeding CatBoost.

### 3.5. Model Interpretability Analysis

To enhance clinical understanding of the prediction mechanism, we applied SHAP analysis to interpret the predictions of the three selected models (Figure 6). The analysis identified the most important features contributing to FC dose prediction in the following order: previous dose, age, and PRC. Specifically, a higher previous dose was associated with a higher predicted FC dose. Conversely, older age or lower PRC levels were linked to a lower predicted FC dose.

**Figure 6.**
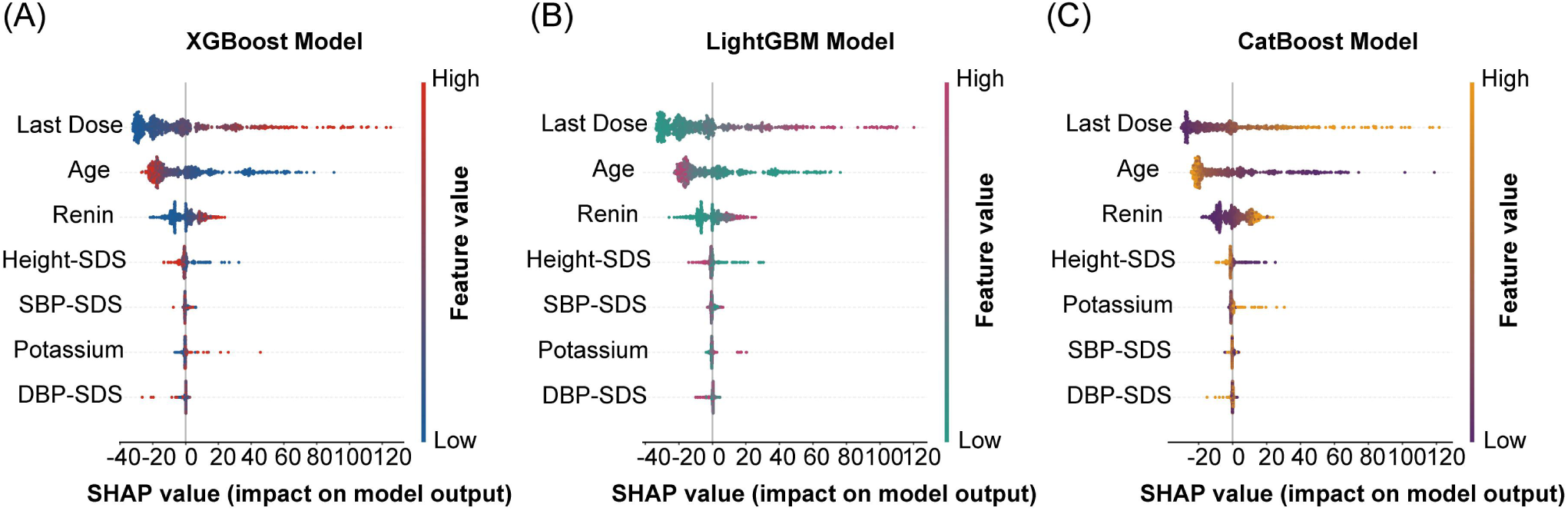
SHAP summary plots for the three final models. **(A)** XGBoost, **(B)** LightGBM, and **(C)** CatBoost. Features are ordered by their mean absolute SHAP value.

SHAP interaction analysis was performed to examine whether the predictive contribution of previous FC dose varied according to concurrent clinical and biochemical indicators (Figure 7). Distinct interaction patterns were observed for all indicators. These patterns indicated that the effect of previous dose on model predictions was not constant across patient states. For example, PRC and serum potassium showed opposite interaction trends between high- and low-level groups as previous dose increased. Overall, these findings suggest that the model incorporated current clinical and biochemical information when using historical dosing information, rather than relying solely on a fixed mapping from previous to current dose.

**Figure 7.**
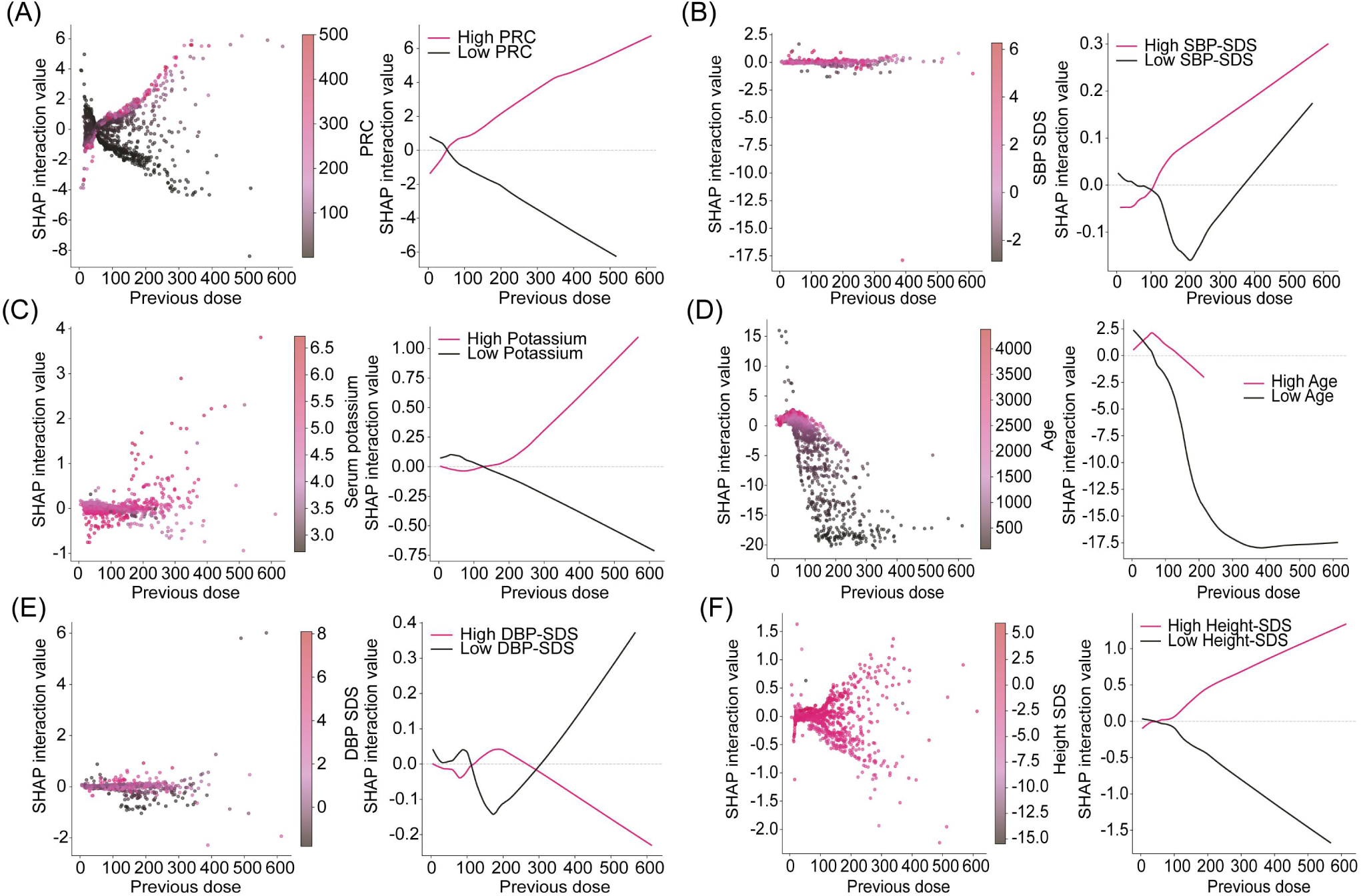
SHAP interaction plots showing the joint effects of previous dose and each covariate on model predictions. For each covariate, the left scatter panel displays individual sample SHAP interaction values coloured by covariate level; the right panel presents LOWESS curves stratified into high and low groups (split by the median value of each covariate). Divergent curve trends between high and low groups indicate strong nonlinear interactions between previous dose and the corresponding covariate. **(A)** Previous Dose x PRC, **(B)** Previous Dose x SBP-SDS, **(C)** Previous Dose x Potassium, **(D)** Previous Dose x Age, **(E)** Previous Dose x DBP-SDS, **(F)** Previous Dose x Height-SDS.

## 4. Discussion

### 4.1. Hypertension Risk Factors

Numerous studies have explored the risk factors for hypertension in patients with CAH, contributing to a systematic understanding of blood pressure regulation in this population. Most findings indicate that lower PRC levels and higher FC doses are associated with elevated blood pressure, although the role of HC remains controversial. Han et al. reported a positive correlation with blood pressure after converting HC doses to prednisone equivalents (Han et al., 2013). In contrast, Maccabee-Ryaboy et al. found no statistically significant association between HC dose and hypertension, regardless of BMI percentile adjustments (Maccabee-Ryaboy et al., 2016). In the present study, we employed multivariable GLMM analysis, with appropriate controls for key confounders, to better delineate the independent effects of medication doses on hypertension risk. Our results identify FC dose as a critical factor influencing hypertension, whereas HC dose has no significant effect.

The pronounced impact of FC on blood pressure aligns with previous research (Verma et al., 2010; Lawrence et al., 2025), underscoring the predominant role of FC excess in hypertension development among CAH patients. This finding also suggests that the elevated DBP-SDS observed in infant patients within our cohort may be attributable to their routine administration of higher FC doses. As a potent mineralocorticoid, FC overdose mimics aldosterone effects, promoting sodium and water retention and blood volume expansion – the core pathophysiological mechanism underlying FC-induced hypertension. Consequently, FC dose titration is crucial for preventing hypertension in CAH patients, particularly those under the age of two.

Our results also revealed a significant inverse association between PRC and hypertension, consistent with the cross-sectional findings of Lubis SM et al., who reported that low PRC levels were linked to hypertension risk (Lubis et al., 2024). Furthermore, the LMM analysis in this study clarified a direct, dose-dependent association between FC dosage and PRC. After adjusting for confounding variables, FC dosage maintained a significant inhibitory effect on PRC. Coupled with the aforementioned significant association between low PRC and an increased risk of hypertension, these findings confirm that PRC may serve as a core monitoring indicator for guiding FC dose adjustments and preventing drug-induced hypertension. Although we observed statistically significant associations between serum sodium/potassium and systolic hypertension, the absolute differences between groups were minimal. Given that sodium and potassium concentrations are influenced by multiple factors, including dietary intake and daily activity, and fluctuate considerably over short periods, the clinical relevance of these statistical differences may be limited. This suggests that serum sodium and potassium may have limited independent interpretive value for hypertension monitoring in CAH and should be considered alongside other indicators for guiding dose adjustments.

Notably, our study found that FC dose was significantly associated with DBP-SDS > 95th percentile, rather than SBP-SDS. Previous work by Neuman showed that DBP-SDS abnormalities persist longer in children than SBP-SDS abnormalities (Neumann et al., 2022). Similarly, Subbarayan et al. employed multiple regression analysis to confirm that FC is a determinant of DBP-SDS but not SBP-SDS (Subbarayan et al., 2014), despite several other studies suggesting that FC affects both systolic and diastolic blood pressure (Bonfig et al., 2016; Tony Nengom et al., 2017). This finding can be explained hemodynamically: FC, by mimicking aldosterone, increases blood volume and peripheral resistance, leading to a more rapid elevation in diastolic pressure. Thus, short-term FC excess may manifest more readily as diastolic hypertension. In contrast, systolic pressure is more substantially influenced by cardiac output and arterial elasticity. It may be more sensitive to long-term vascular structural changes, potentially requiring a longer duration to exhibit significant increases.

In summary, our multivariable analysis highlights the importance of careful FC dose titration, particularly in children aged 0–2 years, whose fluid and electrolyte metabolism is still developing. Monitoring diastolic blood pressure and PRC may serve as sensitive indicators for assessing the risk of FC overdose.

### 4.2. Machine Learning Model Development

Building on the identification of hypertension risk factors, we developed machine learning models to predict follow-up fludrocortisone dosing in patients with CAH, with the aim of supporting individualized longitudinal dose adjustment. The predictor variables included in the feature selection phase are commonly monitored parameters in CAH management. Additionally, SHAP analysis of the optimal models identified the three most important predictive variables: the previous FC dose, age, and PRC.

Age is a crucial reference for FC dose adjustment, with older patients typically being associated with lower predicted FC doses. In contrast, neonates and infants exhibit physiological resistance to mineralocorticoids, which means they require higher FC doses to effectively prevent salt-wasting crises. As children grow and their sensitivity to FC increases, it is essential to gradually reduce the doses to avoid overdose and associated blood pressure abnormalities (Mallappa and Merke, 2022). Meanwhile, although young patients require higher therapeutic doses due to physiological resistance, this also places them at a higher risk of hypertension. In response to this, the SHAP interaction analysis revealed that the model demonstrated a clear protective regulatory mechanism: as the previous dose rose, the negative interaction effect in the low-age group progressively intensified, whereby the model constrained the target FC dose to evade the risk of hypertension in young patients. Conversely, in the high-age group, this interaction effect remained stable near zero, indicating that the model tended to continue with the previous dose. PRC and serum potassium levels reflect the body’s fluid and electrolyte homeostasis. Our models suggest that lower PRC or potassium levels correlate with lower predicted FC doses; SHAP interaction analysis further reveals that under low PRC or low serum potassium conditions, the negative interaction effect significantly intensifies as the previous dose rises. This suggest that FC supplementation should be moderated when these values are low. PRC is particularly sensitive to changes in volume status and is negatively regulated by it; FC overdose leads to volume expansion, suppressing renin secretion. Consequently, PRC is already recognized as a sensitive indicator for adjusting FC doses in clinical practice (Auer et al., 2023). Blood pressure (SBP-SDS and DBP-SDS) serves as the most direct clinical monitoring parameter for assessing the appropriateness of FC dosing. Our model showed that higher BP-SDS values are associated with lower required FC doses. Additionally, Height-SDS emerges as an important variable, with lower Height-SDS associated with higher predicted FC doses. We presume that this relationship does not reflect a direct physiological link between Height-SDS and FC requirement, but rather illustrates a critical trade-off strategy employed by clinicians in practice. Inadequate glucocorticoid (HC) therapy is a common reason for impaired growth in children with CAH. When managing patients with compromised height growth (low Height-SDS), clinicians may reduce HC doses to preserve growth potential, while increasing FC doses to ensure adequate mineralocorticoid activity.

Further analysis of the SHAP interaction plot reveals a downward trend in the previous dosage curve within the low-Height-SDS group. This suggests that if a patient remains at a low Height-SDS despite having previously received high-dose FC therapy, continuing to increase the dose may pose a risk of overdose and induce hypertension. This reflects the model’s risk-aversion mechanism in individualized dosage adjustments. The feature ablation and baseline comparison analysis showed that previous FC dose provided substantial incremental predictive value, while the no-previous-dose model retained meaningful performance. This finding suggests that current clinical and biochemical variables contain predictive information beyond previous dose and may contribute to longitudinal follow-up dose prediction. Currently, adjusting FC dosing for CAH patients primarily relies on dynamically monitoring clinical features and biochemical parameters. The core objective is to prevent salt-wasting while adjusting the dose to avoid hypertension. Substantial evidence demonstrates an inverse correlation between PRC levels and hypertension in CAH patients, establishing PRC as a sensitive indicator for FC dose titration in clinical practice. However, the considerable variability, both inter-individual and intra-individual, in sensitivity to FC complicates the determination of the appropriate dose for specific clinical contexts. Dose adjustments, therefore, depend on dynamic monitoring of these parameters, leading to a reactive approach with time-lagged adjustment.

In this work, we leveraged real-world clinical data to create a high-quality dataset by excluding visits that reflected suboptimal therapy. This strategy allowed the models to learn clinically curated follow-up FC dosing patterns. We then applied advanced machine learning methods to capture complex nonlinear relationships among clinical variables, developing predictive models for FC dose optimization that demonstrated excellent performance on the test set. To our knowledge, this is the first application of machine learning models for medication dose prediction in CAH, aligning with numerous previous studies that indicate that machine learning can enhance prediction accuracy for dosing models of various drugs (e.g., warfarin, tacrolimus) (Asiimwe et al., 2022; Yoon et al., 2024). Notably, our results revealed subtle but significant performance differences among the top three models across age subgroups. This variation could help clinicians choose the most suitable prediction model based on patient age.

### 4.3. Limitations

This study has several inherent limitations. First, because the model incorporates previous FC dose as an essential input, it is designed to support longitudinal dose adjustment rather than initial dose selection. Future studies are warranted to develop complementary models for initial dose determination at diagnosis. The inclusion of previous dose also introduces important interpretational limitations. Previous dose may capture various unmeasured latent factors such as disease severity and adherence, and therefore may be affected by confounding by indication. Therefore, the model outputs should be interpreted only as predictive associations and should not be used for causal inference. Although expert review was performed to exclude visits with clearly inappropriate mineralocorticoid replacement, this process could not establish that all retained dose decisions were optimal. Residual treatment inertia inherent to previous dose may therefore remain. Therefore, the model’s findings should not be overinterpreted as establishing an optimized or causal dosing strategy. Prospective validation is required to determine whether model-assisted dose adjustment improves biochemical control or clinical outcomes beyond usual care. Second, to improve cohort homogeneity, this study applied stringent inclusion and exclusion criteria, resulting in the exclusion of approximately half of the initially screened patients, the majority of whom presented with unstable clinical conditions. They may have reduced the representativeness of the study population and increased the risk of selection bias. Therefore, both the prediction model and the associations estimated in the GLMMs should be interpreted as applying primarily to this selected follow-up cohort. Caution is warranted when interpreting model predictions or implementing this model in routine clinical practice. This selection process may also have led to overoptimistic estimates of model performance. Future prospective multicenter validation in more heterogeneous patient populations is needed. Additionally, missing data handling may have introduced uncertainty. For example, height and weight were interpolated when missing, and although the missing rate for these variables was very low, this procedure assumes that values could be reasonably estimated from adjacent measurements. Non-linear changes in growth or weight would violate this assumption and introduce residual bias. Third, although we observed an inverse association between PRC and hypertension, a methodological limitation of this study is the use of direct PRC rather than plasma renin activity (PRA). PRA has traditionally been widely used for monitoring mineralocorticoid replacement therapy in patients with CAH (Sólyom and Gaál, 1979; Merke and Kabbani, 2001). Although PRC is highly automated and analytically reproducible on modern laboratory platforms (Campbell et al., 2009), it does not account for variation in endogenous substrate availability, particularly angiotensinogen. Therefore, PRC may not fully reflect the functional activity of the renin-angiotensin-aldosterone system. Because this biological variation and potential assay-related measurement error could not be directly accounted for in our multivariable generalized linear mixed models, the estimated regression coefficients for both the association between renin levels and hypertension risk and the association between FC dose and renin levels may have been biased in either direction. Accordingly, these effect estimates should be interpreted with caution. Fourth, the substantial missingness of certain variables (e.g., testosterone and androstenedione) prevented their inclusion in the multivariable analysis and model development. Future studies using more complete datasets are necessary to corroborate our findings and refine the models. Furthermore, successfully integrating the predictive models into clinical workflows is crucial; therefore, we plan to develop user-friendly interactive interfaces to enhance the clinical utility and adoption of these tools.

### 4.4. Conclusion

In summary, our study confirms the central role of FC dose in the development of hypertension among CAH patients. More importantly, it demonstrates the feasibility of using machine learning models as decision-support tools for individualized follow-up FC dose adjustment. This achievement outlines a viable pathway toward personalized dosing in the management of rare diseases. Looking ahead, with multi-center validation and clinical deployment of these models, replacement therapy for CAH holds the potential to transition from empirical decision-making to data-driven, precise regulation.

## Data Availability

All data produced in the present study are available upon reasonable request to the authors

## Acknowledgements

We would like to thank Professor Shenglong Li for his invaluable advice and sustained mentorship, and Teng Wang for his expert input on the methodological aspects. We also extend our sincere appreciation to Zhixin Luo, Zhenyang Dai, Yujie Luo, Yi Yang, Junhao He, Guanxi Zhu and Song Jiang for their technical assistance with this project.

