## Supplemental Materials for "Precision Management of Fludrocortisone-Related Hypertension Risk in Congenital Adrenal Hyperplasia: A Machine Learning Approach to Personalized Dosing"

**Figure S1.** The black solid line represents the point estimate of the FC dose coefficient, and the gray band indicates its 95% CI across varying delta levels (Delta = ±0.1, ±0.3, ±0.5 SD).

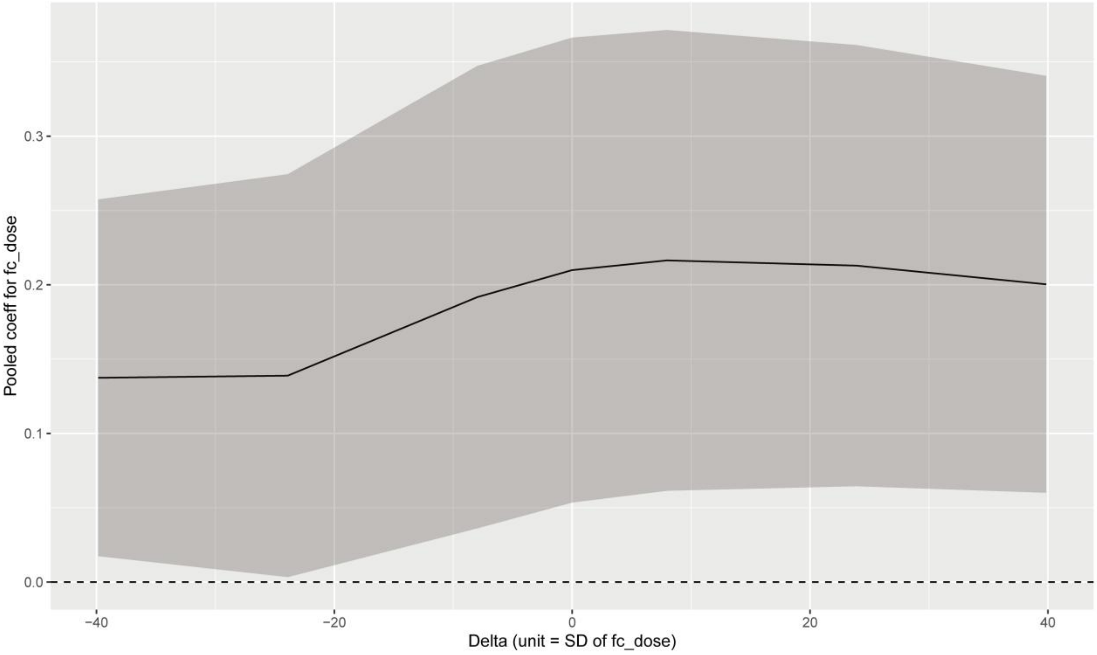


**Figure S2.** The black solid line represents the point estimate of the FC dose coefficient, and the gray band indicates its 95% CI across varying delta levels (Delta = ±0.1, ±0.3, ±0.5 SD).

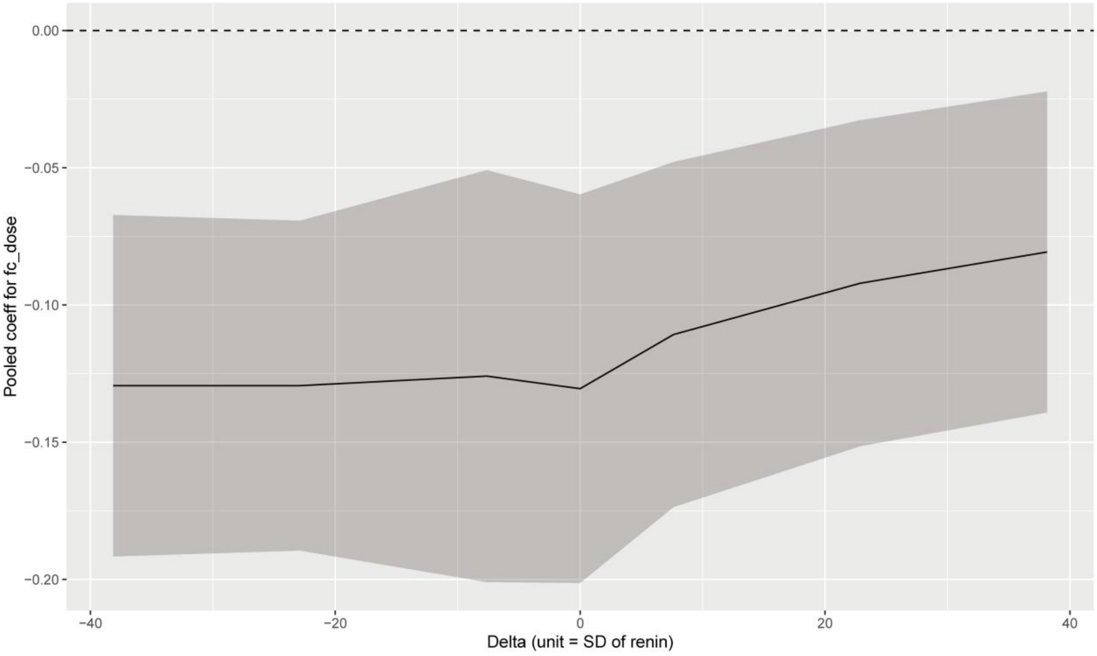


**Table S1.** Performance comparison among the full-feature, no-previous-dose, and previous-dose-only models.

| Model | $R^2$ | MAE | RMSE | MAPE |
|---|---|---|---|---|
| Full Model | 0.77 | 17.88 | 29.25 | 0.33 |
| no-previous-dose Model | 0.59 | 26.64 | 40.73 | 0.47 |
| previous-dose-only Model | 0.56 | 25.12 | 42.65 | 0.48 |

**Table S2.** Predictive Performance of Machine Learning Models Across Age Subgroups.

| Subgroups Based on Age | | | |
|---|---|---|---|
| | Under (%) | Ideal (%) | Over (%) |
| 0-2 year (n = 209) | | | |
| CatBoost | 22.1 (15.5-26.8) | 60.6 (54.6-67.5) | 17.3 (12.2-23.5) |
| XGBoost | 17.7 (14.1-22.3) | 60.1 (55.3-64.1) | 22.2 (19.8-25.6) |
| LightGBM | 24.9 (18.4-30.6) | 58.0 (51.4-64.4) | 17.1 (12.9-23.0) |
| 2-8 years (n = 418) | | | |
| CatBoost | 11.7 (9.0-14.9) | 53.2 (49.0-57.2) | 35.1 (30.6-39.1) |
| XGBoost | 10.7 (8.2-13.8) | 53.4 (49.0-57.9) | 36.0 (31.2-39.5) |
| LightGBM | 11.1 (8.4-14.6) | 54.9 (50.7-59.1) | 34.0 (29.2-37.5) |
| 8-12 years (n = 114) | | | |
| CatBoost | 11.3 (4.8-17.4) | 38.9 (32.2-47.0) | 49.8 (42.2-58.3) |
| XGBoost | 10.4 (3.9-16.1) | 45.0 (36.5-52.6) | 44.6 (37.4-53.9) |
| LightGBM | 11.3 (4.8-17.9) | 45.7 (37.4-37.4-53.9) | 43.0 (35.2-52.2) |

Predictions were categorized as **Under** (<80% of actual dose), **Ideal** (within 20% of actual dose, i.e., 80–120%), or **Over** (>120% of actual dose). Values are percentages (95% confidence intervals).

**Table S3.** Bonferroni-Adjusted P-Values for Pairwise Comparisons of Model Performance Across Age Subgroups.

| Model | Model2 | 0-2 years | 2-8 years | 8-12 years |
|---|---|---|---|---|
| CatBoost | XGBoost | 0.01 | 0.8 | $2.3x10^{-17}$ |
| XGBoost | LightGBM | 0.09 | $8.7x10^{-6}$ | 0.6 |
| CatBoost | LightGBM | $3.0x10^{-6}$ | $7.9x10^{-6}$ | $6.9x10^{-19}$ |

**Table S4.** Bonferroni-Adjusted P-Values for Comparisons of Ideal Prediction Rate (Withing 20% of actual) Across Age Subgroups

| Model | Subgroup1 | Subgroup2 | P value |
|---|---|---|---|
| CatBoost | 0-2 years | 2-8 years | $8.2x10^{-29}$ |
| | 2-8 years | 8-12 years | $7.1x10^{-34}$ |
| | 0-2 years | 8-12 years | $8.6x10^{-34}$ |
| XGBoost | 0-2 years | 2-8 years | $7.8x10^{-25}$ |

| | | | |
|---|---|---|---|
| | 2-8 years | 8-12 years | $3.3 \times 10^{-33}$ |
| | 0-2 years | 8-12 years | $3.2 \times 10^{-28}$ |
| LightGBM | 0-2 years | 2-8 years | $8.8 \times 10^{-11}$ |
| | 2-8 years | 8-12 years | $3.5 \times 10^{-32}$ |
| | 0-2 years | 8-12 years | $3.0 \times 10^{-30}$ |
